# Obesity-Associated Genetic Variants and AMPK Signaling in Cardiovascular Disease: A Systematic Review of Mechanisms and Clinical Implications

**DOI:** 10.1101/2025.10.04.25337310

**Authors:** Ifrah Siddiqui, Nabeel Ahmad Khan, Zille Haider

## Abstract

**Objective:** To investigate how genetic variations in obesity-related genes affect the AMPK signaling pathway and contribute to the pathophysiology of cardiovascular disease in the context of obesity.

**Background:** The global rise in obesity presents serious public health challenges, significantly increasing the risk of comorbidities such as type 2 diabetes and cardiovascular disease. Genetic predisposition plays a critical role in obesity, with several genes influencing metabolic regulation. AMP-activated protein kinase (AMPK) is a central regulator of cellular energy homeostasis. Disruptions in its signaling pathway may contribute to metabolic dysfunction and cardiovascular complications. Understanding how genetic variations impact AMPK signaling is essential for the development of targeted interventions.

**Methods:** A systematic literature review was conducted using databases including PubMed, MEDLINE, Google Scholar, and both open-access and subscription-based journals. The search focused on studies examining genetic variations in obesity-associated genes—FTO, MC4R, LEP, LEPR, PCSK1, PPARG, BDNF, SIM1, TBC1D1, ADRB3, UCP1, and SH2B1—and their impact on AMPK signaling. No date restrictions were applied. Articles were selected based on predefined inclusion criteria and assessed in accordance with PRISMA guidelines to evaluate the mechanisms linking genetic variants with AMPK modulation, energy balance, inflammation, and cardiovascular risk.

**Results:** Genetic variants in the selected obesity-related genes were found to significantly influence AMPK activation. These alterations led to impaired energy expenditure, increased lipid storage, and disturbances in glucose metabolism. Dysregulation of AMPK signaling also promoted chronic low-grade inflammation and endothelial dysfunction—factors closely associated with elevated cardiovascular disease risk. The findings underscore the pivotal role of gene-AMPK interactions in metabolic and cardiovascular health.

**Conclusion:** Genetic variations in obesity-related genes impair AMPK activation, disrupting energy metabolism and promoting inflammation and vascular dysfunction. This contributes to increased cardiovascular risk in obese individuals. Personalized therapies targeting AMPK activation and gene-specific pathways may offer promising strategies to counteract obesity-induced metabolic dysregulation and improve cardiovascular outcomes.

## Background

The global prevalence of obesity has reached epidemic proportions, with significant implications for public health. Obesity is a complex, multifactorial condition that increases the risk of various comorbidities, including type 2 diabetes, hypertension, and cardiovascular disease. Understanding the underlying mechanisms that contribute to obesity is critical for developing effective prevention and intervention strategies [1, 2].

Genetic factors play a substantial role in the susceptibility to obesity, influencing individual responses to environmental stimuli such as diet and physical activity. Recent advances in genomic research have identified several obesity-related genes, including *FTO*, *MC4R*, *LEP*, *LEPR*, *PCSK1*, *PPARG*, *BDNF*, *SIM1*, *TBC1D1*, *ADRB3*, *UCP1*, and *SH2B1*. These genes are involved in various metabolic processes, including appetite regulation, energy expenditure, and insulin sensitivity. However, the mechanisms by which genetic variations in these genes affect metabolic pathways, particularly the AMPK (AMP-activated protein kinase) signaling pathway, remain poorly understood [2, 3].

AMPK is a central regulator of energy homeostasis, coordinating cellular responses to metabolic stress. Dysregulation of AMPK signaling has been linked to obesity and related metabolic disorders, contributing to inflammation, oxidative stress, and cardiovascular pathology. This study aims to explore how genetic variations in obesity-related genes influence AMPK activation and its downstream effects, particularly in the context of cardiovascular disease [3, 4].

By investigating these relationships, this research addresses a critical gap in our understanding of the interplay between genetics, metabolism, and cardiovascular health. The findings have the potential to inform personalized approaches to obesity management and cardiovascular disease prevention, ultimately contributing to improved health outcomes for affected individuals and populations [4, 5].

## Methods

### Aim of the Study

This study aims to investigate how genetic variations in specific obesity-related genes—FTO, MC4R, LEP, LEPR, PCSK1, PPARG, BDNF, SIM1, TBC1D1, ADRB3, UCP1, and SH2B1—influence the AMPK (AMP-activated protein kinase) signaling pathway. The goal is to explore the molecular mechanisms by which these variations impact AMPK activation and its downstream targets, and to assess how altered AMPK signaling contributes to energy dysregulation, inflammation, and cardiovascular disease (CVD) within the context of obesity. The study also aims to evaluate how these gene–pathway interactions serve as potential mediators between genetic obesity risk and cardiovascular pathology.

### Research Question

How do genetic variations in key obesity-related genes impact AMPK signaling and its downstream metabolic and inflammatory pathways, and how does this disruption contribute to the pathogenesis of cardiovascular disease in individuals with obesity?

### Search Focus

A comprehensive and systematic literature review was conducted using the PUBMED, MEDLINE, and Google Scholar databases, along with access to relevant open-access and subscription-based journals. No publication date restrictions were applied.

The search focused on studies examining:

- Genetic polymorphisms or mutations in obesity-related genes: FTO, MC4R, LEP, LEPR, PCSK1, PPARG, BDNF, SIM1, TBC1D1, ADRB3, UCP1, and SH2B1
- AMPK pathway regulation and signaling mechanisms
- The impact of gene-AMPK interactions on metabolic homeostasis, lipid metabolism, glucose regulation, inflammation, and cardiovascular outcomes

– The mechanistic link between obesity-driven genetic variations, AMPK dysregulation, and CVD development The literature was screened and selected based on relevance to the study’s objectives. Literature search began in August 2021 and ended in March 2025. An in-depth investigation was conducted during this duration based on the parameters of the study as defined above. During revision, further literature was searched and referenced until September 2025. The literature search and all sections of the manuscript were checked multiple times during the months of revision (April 2025 September 2025) to maintain the highest accuracy possible. This approach allowed for a comprehensive investigation of the impact of genetic variations in obesity-related genes on AMPK signaling and cardiovascular risk, adhering to the PRISMA (Preferred Reporting Items for Systematic Reviews and Meta-Analyses) guidelines for systematic reviews.

### Search Queries/Keywords

**General Search Terms:**

- “Obesity genes” OR “genetic obesity” OR “obesity-related polymorphisms”
- “AMPK signaling” OR “AMPK activation” OR “AMPK dysregulation”
- “Obesity AND cardiovascular disease”
- “AMPK pathway AND cardiovascular outcomes”
- “Obesity AND inflammation AND AMPK”

**Gene-Specific Searches:**

- “FTO polymorphism” AND “AMPK signaling”
- “MC4R gene variant” AND “obesity and AMPK”
- “LEP mutation” OR “leptin gene” AND “AMPK pathway”
- “LEPR variants” AND “AMPK activation”
- “PCSK1 mutation” AND “AMPK”
- “PPARG AND AMPK signaling AND lipid metabolism”
- “BDNF AND obesity AND AMPK”
- “SIM1 gene AND AMPK”
- “TBC1D1 AND glucose metabolism AND AMPK”
- “ADRB3 gene” AND “thermogenesis AND AMPK”
- “UCP1 polymorphism” AND “energy expenditure AND AMPK”
- “SH2B1 AND AMPK AND insulin sensitivity”

**Pathway and Mechanism Specific Terms:**

- “AMPK and energy homeostasis”
- “AMPK and inflammation”
- “AMPK and cardiovascular disease”
- “Genetic regulation of AMPK”
- “Obesity genes and metabolic pathways”

Boolean operators (AND, OR) were used to construct targeted search queries to retrieve literature focusing on the molecular and pathophysiological relationships between obesity-related genes and AMPK signaling.

### Objectives of the Search

- To understand how genetic variations in obesity-associated genes alter AMPK activity
- To explore the downstream metabolic and inflammatory effects of these alterations
- To assess how AMPK-mediated dysregulation contributes to the development of cardiovascular diseases in obese individuals

### Screening and Eligibility Criteria

#### Initial Screening

Titles and abstracts were screened for relevance to AMPK signaling, obesity gene variants, and cardiovascular consequences.

#### Full-Text Review

Articles meeting initial criteria were reviewed in full. Studies were selected if they provided mechanistic or clinical insight into the interplay between obesity-related genes and AMPK signaling, especially in the context of metabolic and cardiovascular consequences.

#### Data Extraction

Key findings were extracted, focusing on gene-AMPK interactions, affected downstream signaling, and cardiovascular or metabolic outcomes.

### Inclusion and Exclusion

**Criteria Inclusion Criteria:**

1. Studies investigating the listed genes (*FTO*, *MC4R*, etc.) and their influence on AMPK pathway
2. Mechanistic studies (in vitro, in vivo, or computational) exploring AMPK-dependent metabolic and inflammatory regulation
3. Studies connecting AMPK dysregulation to cardiovascular pathology in the context of obesity

**Exclusion Criteria:**

1. Studies lacking investigation into either the AMPK pathway or gene-specific impacts
2. Reviews or studies focusing only on general obesity without reference to genetic mechanisms or AMPK
3. Low methodological quality: inadequate controls, poorly defined endpoints, or lack of reproducibility
4. Articles not in English
5. Non-peer-reviewed sources (e.g., conference abstracts, editorials, theses)
6. Duplicate publications or redundant datasets
7. Studies not relevant to cardiovascular outcomes or systemic effects of AMPK dysregulation

**Rationale for Screening and Inclusion**

- **FTO:** Alters energy intake and expenditure; linked to AMPK suppression in hypothalamic regions and adipocytes.
- **MC4R:** Affects appetite regulation; modulates AMPK signaling in the hypothalamus.
- **LEP & LEPR:** Leptin signaling activates AMPK in the hypothalamus and peripheral tissues; resistance disrupts AMPK-mediated energy sensing.
- **PCSK1:** Influences prohormone processing affecting AMPK-regulated appetite and energy metabolism.
- **PPARG:** Regulates adipogenesis and lipid metabolism; interacts with AMPK in insulin signaling.
- **BDNF & SIM1:** Central regulators of energy homeostasis; modulate AMPK activity through hypothalamic pathways.
- **TBC1D1:** Directly interacts with AMPK; critical in skeletal muscle glucose uptake.
- **ADRB3:** Influences thermogenesis and lipolysis via AMPK-related pathways.
- **UCP1:** Thermogenic gene regulated via AMPK-activated PGC-1α.
- **SH2B1:** Enhances insulin and leptin signaling; affects AMPK activation and glucose metabolism.

These genes play diverse roles across central and peripheral tissues, and their variants can modulate AMPK signaling cascades that influence metabolic flexibility, inflammation, and cardiovascular health.

### Assessment of Article Quality and Potential Biases

- **Quality Assessment:** Each article was evaluated for methodological rigor, including use of appropriate controls, clear mechanistic endpoints, and reproducibility. Peer-reviewed status was prioritized to ensure scientific integrity.
- **Bias Assessment:**

**Publication Bias:** To counteract positive-result bias, broad searches included both favorable and unfavorable findings regarding gene–AMPK interactions.
**Selection Bias: Predefined** inclusion/exclusion criteria were rigorously applied.
**Reporting Bias: Articles** were examined for inconsistencies or lack of data; all included papers underwent multiple rounds of full-text review.

### Language and Publication Restrictions

Only English-language, peer-reviewed articles were included. There were no date restrictions to allow inclusion of both foundational studies and recent advances. Unpublished manuscripts and grey literature were excluded to maintain a high standard of scientific evidence.

### Results

A total of 2020 articles were identified using database searching, and 1925 were recorded after duplicates removal. 1638 were excluded after screening of title/abstract, 161 were finally excluded, and 6 articles were excluded during data extraction. These exclusions were primarily due to factors such as non-conformity with the study focus, insufficient methodological rigor, or data that did not align with the research questions. Finally, 120 articles were included as references. PRISMA Flow Diagram is Fig 1.

**Fig 1.**
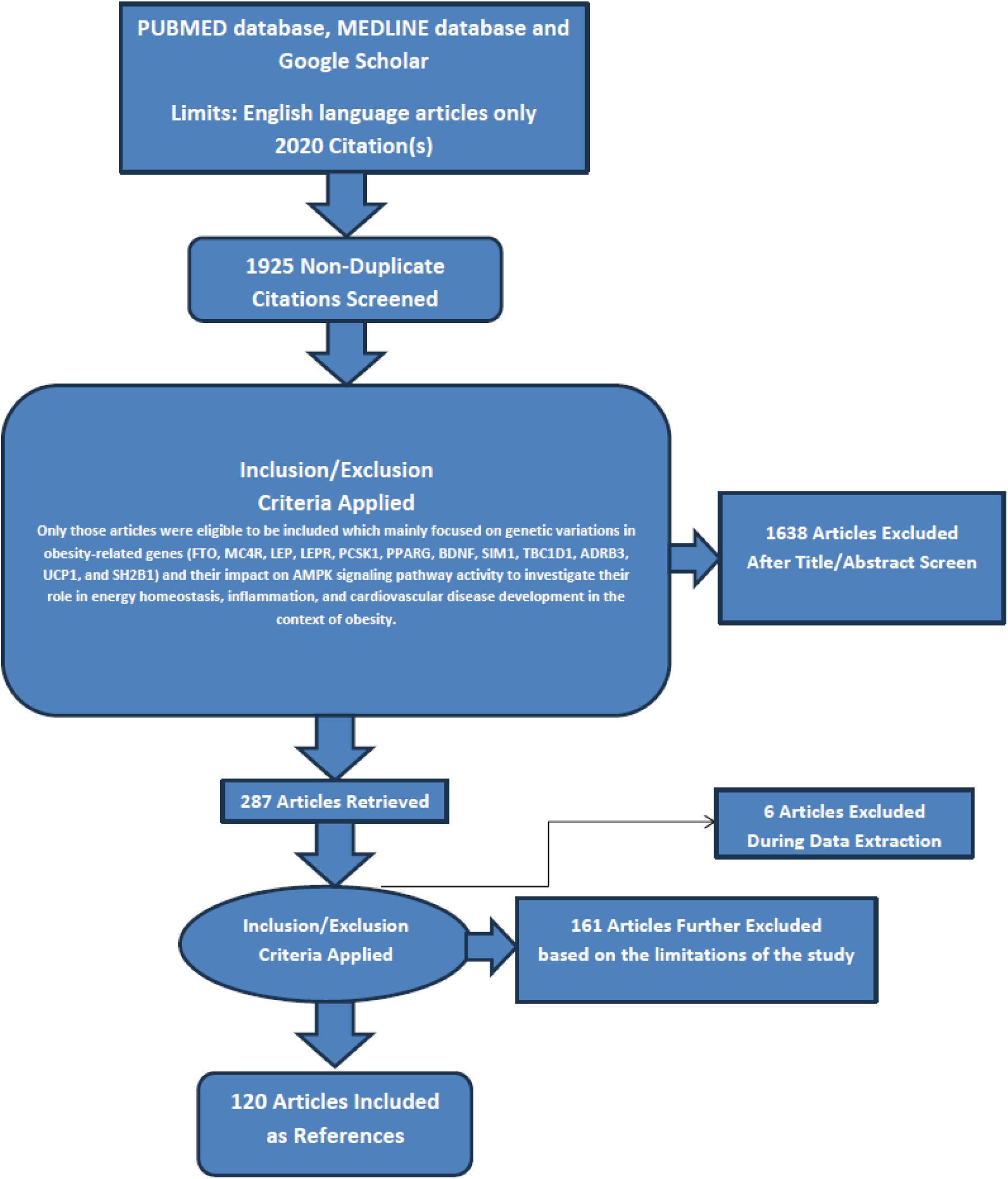
PRISMA FLOW DIAGRAM: This figure represents graphically the flow of citations in the study.

### The Role of Obesity-Linked Gene Variants in AMPK Activation and Cardiovascular Disease

1. FTO

**Genetic Architecture of FTO and Its Metabolic Influence:**
Genetic variations in the FTO (Fat Mass and Obesity-Associated) gene have consistently been linked to increased susceptibility to obesity. Among these, the single nucleotide polymorphism (SNP) rs9939609 is one of the most extensively studied. Individuals carrying the risk allele exhibit increased body mass index (BMI) and a higher prevalence of obesity. The biological consequences of this SNP appear to involve dysregulation of energy metabolism pathways, particularly those governed by AMP-activated protein kinase (AMPK) [5, 6].
AMPK serves as a central metabolic sensor that maintains cellular energy homeostasis. Variants in FTO may modulate AMPK activation indirectly by influencing adipogenesis and lipid metabolism. Accumulation of intracellular lipids, especially free fatty acids, alters cellular AMP/ATP ratios, potentially activating AMPK under some metabolic stress conditions. However, overexpression of FTO has also been associated with suppressed AMPK phosphorylation, leading to impaired AMPK activation. This suppression is likely mediated through enhanced ATP-consuming anabolic processes, driven by increased expression of adipogenic genes and alterations in adipocyte differentiation [6, 7].
**Mechanistic Pathways Linking FTO to AMPK Dysregulation:**
Variants in FTO impact lipid and glucose metabolism in a manner that disrupts AMPK’s regulatory role in energy balance. Reduced AMPK activity contributes to enhanced lipogenesis and diminished fatty acid oxidation, promoting lipid accumulation and increasing insulin resistance. Impaired AMPK function is also associated with reduced translocation of GLUT4 transporters to the plasma membrane, limiting glucose uptake and exacerbating hyperglycemia. These metabolic disturbances contribute to the development of metabolic syndrome and increase cardiovascular risk [7, 8].
AMPK negatively regulates the mechanistic target of rapamycin (mTOR) pathway. In FTO-variant carriers, decreased AMPK activity may result in unchecked mTOR signaling, leading to enhanced protein synthesis, adipogenesis, and chronic inflammation. Increased mTOR activity is implicated in endothelial dysfunction and atherosclerotic progression. Additionally, FTO is highly expressed in the hypothalamus, where it may alter AMPK-mediated appetite regulation. Disruption in hypothalamic AMPK signaling can lead to increased caloric intake and reduced energy expenditure, compounding the risk of obesity [8, 9].
**Cardiovascular Effects Mediated by Altered AMPK Signaling:**
Disruption of AMPK signaling due to FTO variants contributes to an energy imbalance characterized by excessive lipid storage and impaired lipid oxidation. The resulting obesity increases cardiovascular disease (CVD) risk through multiple mechanisms, including elevated blood pressure, dyslipidemia, and insulin resistance [9].
Reduced AMPK activity also promotes pro-inflammatory states via diminished inhibition of the NF-κB pathway, a key driver of vascular inflammation. Chronic low-grade inflammation is a central factor in the initiation and progression of atherosclerosis. Endothelial dysfunction is further exacerbated by insufficient AMPK-mediated nitric oxide (NO) production and elevated oxidative stress, both of which facilitate plaque formation and vascular damage [9, 10].
In the myocardium, AMPK supports energy production and contractile function. Impairments in this pathway can disrupt myocardial metabolism, contributing to cardiac dysfunction. In individuals with FTO-associated obesity, these molecular changes can worsen hypertension and precipitate heart failure [10].
**Clinical Relevance:**
Understanding how FTO variants disrupt AMPK signaling offers a mechanistic explanation for the elevated cardiovascular risk observed in obese individuals with these genetic polymorphisms. This insight is particularly important in stratifying patients based on genetic risk and tailoring therapeutic interventions accordingly. The convergence of metabolic dysfunction, inflammation, and vascular injury underscores the clinical importance of early identification and management of FTO-mediated metabolic derangements [5].
**Therapeutic Implications:**
Interventions targeting AMPK activation represent a promising strategy for mitigating the adverse cardiovascular effects associated with FTO variants. Pharmacological agents such as metformin and AICAR, which activate AMPK, may help restore metabolic balance, reduce inflammation, and improve endothelial function in at-risk populations. These agents may also attenuate the downstream activation of the mTOR pathway, offering additional cardiometabolic benefits [6].
Physical activity serves as a physiological AMPK activator, suggesting that lifestyle modification may offer non-pharmacological means of counteracting the metabolic effects of FTO polymorphisms. Regular aerobic exercise improves insulin sensitivity, promotes lipid oxidation, and supports cardiovascular health through AMPK-dependent mechanisms [7, 8].
Future therapeutic strategies may involve precision medicine approaches aimed at modulating the interaction between FTO and AMPK. Such targeted interventions could offer improved outcomes in managing obesity-related cardiovascular complications [8].
**Implications:**
This study highlights the potential for incorporating genetic screening into cardiovascular risk assessment, particularly in obese individuals. Identification of FTO risk alleles may guide early preventive strategies and inform personalized treatment plans. The link between FTO, AMPK, and cardiovascular health also supports the development of novel therapeutics targeting this molecular axis. By addressing the upstream genetic and metabolic dysregulation, such interventions could offer more durable and effective cardiovascular risk reduction [9].
Genetic variants in the FTO gene contribute to the dysregulation of AMPK signaling, promoting obesity and increasing susceptibility to cardiovascular disease. These variants impair energy homeostasis, promote inflammation, and disrupt vascular function. Targeting the AMPK pathway through pharmacologic and lifestyle interventions presents a viable strategy for managing the cardiometabolic consequences of FTO-associated obesity. Understanding the mechanistic interplay between FTO and AMPK offers critical insights into the pathogenesis and therapeutic opportunities for obesity-related cardiovascular disorders [9, 10].
2. MC4R

**MC4R Variants and Energy Regulation:**
Genetic variations in the Melanocortin-4 Receptor (MC4R) gene are strongly associated with increased susceptibility to obesity. The MC4R receptor is a critical regulator of energy balance, appetite, and metabolic homeostasis. Variants such as rs17782313, located near the MC4R locus, have been linked to elevated body mass index (BMI) and increased food intake. These polymorphisms may alter receptor function or expression, disrupting neural circuits involved in feeding behavior and energy expenditure [11].
MC4R is prominently expressed in the hypothalamus, where it modulates appetite and sympathetic nervous system (SNS) activity. Disruption of this receptor’s function by genetic variants leads to dysregulation of downstream metabolic pathways, including AMP-activated protein kinase (AMPK) signaling. AMPK serves as a cellular energy sensor and regulator, and its dysfunction plays a key role in the pathogenesis of obesity-related metabolic disorders [11, 12].
**Disruption of AMPK Signaling by MC4R Variants:**
The MC4R pathway intersects with AMPK signaling in both central and peripheral tissues. In the hypothalamus, MC4R activation normally suppresses AMPK activity to promote satiety. Variants that impair MC4R function reduce this suppression, resulting in increased hypothalamic AMPK activation, which may paradoxically stimulate appetite and food intake. This altered regulatory loop contributes to positive energy balance and obesity development [12].
Peripheral metabolic consequences of MC4R dysfunction include reduced AMPK activation in tissues such as liver and skeletal muscle. This results from energy excess due to increased caloric intake, leading to diminished fatty acid oxidation, impaired glucose uptake, and enhanced lipid storage. These changes foster systemic insulin resistance and metabolic inflexibility [12, 13].
MC4R also influences SNS output, which affects thermogenesis and energy expenditure. Reduced SNS activity in individuals with MC4R variants may blunt AMPK-driven thermogenic responses, favoring energy conservation and fat accumulation. These physiological alterations extend the metabolic impact of MC4R polymorphisms beyond central appetite regulation [13].
**Hormonal Crosstalk and Metabolic Dysregulation:**
The interaction between MC4R and metabolic hormones such as leptin and insulin is critical to energy homeostasis. Leptin normally acts on the hypothalamus to suppress hunger and inhibit AMPK activity. However, MC4R variants are associated with leptin resistance, characterized by diminished central leptin responsiveness and compensatory activation of hypothalamic AMPK. This state reinforces hyperphagia and obesity [13, 14].
Similarly, MC4R dysfunction is linked to insulin resistance. In peripheral tissues, insulin resistance impairs AMPK activation, compromising cellular glucose uptake and promoting lipid accumulation. These combined effects further entrench metabolic dysfunction and increase the risk of obesity-related complications [14].
**Cardiovascular Consequences of AMPK Dysregulation:**
Variants in MC4R contribute to cardiovascular disease (CVD) risk through their impact on AMPK-mediated metabolic control. Chronic energy imbalance driven by increased food intake and reduced energy expenditure promotes obesity, a major CVD risk factor. In obese individuals with impaired AMPK signaling, metabolic disturbances such as dyslipidemia, hypertension, and glucose intolerance are more pronounced [14, 15].
AMPK possesses anti-inflammatory properties through inhibition of NF-κB and other inflammatory mediators. Disruption of AMPK activation due to MC4R variants may exacerbate systemic inflammation, accelerating atherosclerosis and vascular injury. Reduced AMPK activity also impairs endothelial nitric oxide (NO) production and increases oxidative stress, leading to endothelial dysfunction—a precursor to plaque formation and cardiovascular events [15].
Hypertension, often present in MC4R-related obesity, may be compounded by altered AMPK signaling that affects vascular tone and cardiac energetics. Collectively, these processes underscore the mechanistic link between MC4R variants, AMPK disruption, and increased cardiovascular morbidity [15, 16].
**Clinical Relevance:**
Understanding the interplay between MC4R genetic variants and AMPK signaling provides critical insight into the molecular basis of obesity and its cardiovascular consequences. Given the prevalence of MC4R-associated obesity, identifying affected individuals through genetic screening could facilitate early intervention strategies. The central and peripheral effects of MC4R variants highlight the need for therapies that address both appetite dysregulation and metabolic inefficiency [16].
AMPK’s role in modulating inflammation, lipid metabolism, and vascular health makes it a compelling target for mitigating cardiovascular risk in this population. Therapeutic approaches aimed at restoring AMPK function may offer cardioprotective benefits in individuals with MC4R-driven obesity [16, 17].
**Therapeutic Interventions and Targeted Strategies:**
Pharmacological activation of AMPK is a promising strategy to counteract the adverse effects of MC4R variants. Agents such as metformin, widely used in type 2 diabetes, activate AMPK and improve metabolic outcomes. Natural compounds like resveratrol and berberine also stimulate AMPK and may provide adjunctive benefits in managing obesity-related metabolic dysfunction [17].
Lifestyle interventions remain essential. Regular physical activity robustly activates AMPK in skeletal muscle and improves insulin sensitivity. Exercise also enhances mitochondrial function and energy expenditure, helping to offset the energy surplus caused by impaired MC4R signaling [17, 18].
Therapies targeting the melanocortin pathway represent an emerging avenue. MC4R agonists or modulators may restore receptor function and normalize appetite signaling. Such treatments could work synergistically with AMPK activators to correct both central and peripheral metabolic imbalances [18].
**Implications:**
The findings underscore the importance of integrating genetic and molecular data into clinical practice. MC4R variants represent a critical node in the regulatory network linking appetite control, energy balance, and cardiovascular health. Personalized interventions based on MC4R genotype and AMPK activity may enhance obesity treatment outcomes and reduce long-term cardiovascular risk [18, 19].
This research supports the development of targeted therapies that bridge neuroendocrine and metabolic regulation. By addressing the root molecular disruptions, such interventions offer a precision medicine approach to managing obesity and its cardiovascular complications [19].
Genetic variations in the MC4R gene contribute to altered AMPK signaling, promoting obesity and increasing cardiovascular disease risk. These variants disrupt central appetite regulation and peripheral energy metabolism, leading to systemic metabolic dysfunction. Targeting AMPK activation and restoring MC4R signaling represent promising strategies for reducing cardiovascular morbidity in genetically predisposed individuals. Understanding these interactions enhances our capacity to develop effective, individualized therapies for obesity-related cardiovascular disease [19, 20].
3. LEP

**Genetic Variants in LEP and Their Effect on Leptin Function:**
The LEP gene encodes leptin, a hormone critical for the regulation of energy homeostasis, appetite control, and energy expenditure. Genetic variants within LEP, such as rs7799039, have been linked to altered leptin production and secretion. These variants are associated with the development of leptin resistance—a condition characterized by elevated circulating leptin levels but diminished physiological response—commonly observed in individuals with obesity [21].
Leptin exerts its metabolic effects through several signaling pathways, including the AMP-activated protein kinase (AMPK) pathway, which acts as a master regulator of cellular energy status. Altered leptin signaling due to LEP variants can disrupt AMPK activation in both central and peripheral tissues, impairing metabolic homeostasis and promoting the pathogenesis of obesity and related cardiovascular disease [21, 22].
**Effects of Leptin Variants on AMPK Signaling:**
In the hypothalamus, leptin normally inhibits AMPK activity to suppress appetite and increase energy expenditure. In the presence of LEP variants that reduce leptin efficacy, this inhibitory effect is diminished. The resulting overactivation of hypothalamic AMPK has been linked to increased food intake and positive energy balance, contributing to weight gain [22].
In peripheral tissues such as skeletal muscle and liver, leptin activates AMPK to promote fatty acid oxidation and enhance glucose uptake. LEP variants that impair leptin production or receptor sensitivity reduce AMPK activation in these tissues, favoring lipid accumulation, impaired glucose metabolism, and decreased energy expenditure. These disruptions promote the development of insulin resistance and exacerbate obesity-related metabolic dysfunction [22, 23].
**Mechanisms of AMPK Dysregulation Linked to LEP Variants:**
Leptin resistance, often arising from LEP polymorphisms, underlies the dysregulation of AMPK activity in multiple organ systems. In skeletal muscle and liver, reduced leptin responsiveness leads to insufficient AMPK activation, suppressing fatty acid oxidation while promoting lipogenesis. The resulting metabolic imbalance contributes to ectopic fat deposition, particularly hepatic steatosis, and systemic insulin resistance [23].
Within the central nervous system, impaired leptin signaling reduces the suppression of hypothalamic AMPK, disrupting appetite control and reinforcing overeating. This feedback loop exacerbates energy imbalance and body weight gain, reinforcing the adverse metabolic consequences of LEP gene variants [23, 24].
Leptin also influences inflammatory pathways by modulating immune cell activity. Variants that lead to high leptin levels or leptin resistance can stimulate pro-inflammatory cytokine release and immune cell activation. AMPK, known for its anti-inflammatory role, normally inhibits nuclear factor-kappa B (NF-κB) signaling. Impaired leptin-mediated AMPK activation may diminish this anti-inflammatory control, contributing to chronic low-grade inflammation—a recognized driver of cardiovascular pathology [24].
**Impact on Cardiovascular Disease via Altered AMPK Signaling:**
Obesity associated with LEP gene variants increases the risk of cardiovascular disease through multiple mechanisms linked to impaired AMPK activity. Reduced AMPK activation in the context of leptin resistance intensifies insulin resistance and disrupts glucose handling, promoting endothelial dysfunction. These metabolic abnormalities contribute to atherosclerosis and elevate the risk of ischemic events [24, 25].
Chronic inflammation, mediated in part by elevated leptin levels and reduced AMPK activity, accelerates vascular damage and plaque formation. Leptin-induced activation of macrophages and pro-inflammatory cytokine production promotes vascular inflammation, a key contributor to the development of coronary artery disease [25].
AMPK supports vascular function by enhancing nitric oxide (NO) production and reducing oxidative stress. When AMPK activation is compromised due to defective leptin signaling, endothelial dysfunction ensues, promoting vascular stiffness and plaque vulnerability. Additionally, leptin’s effect on the sympathetic nervous system may elevate blood pressure. Variants that increase leptin levels can exacerbate sympathetic overactivity, contributing to hypertension—a major modifiable risk factor for cardiovascular events [25, 26].
**Clinical Relevance:**
The interaction between LEP gene variants and AMPK signaling provides insight into the molecular mechanisms linking obesity with cardiovascular disease. Individuals harboring pathogenic LEP polymorphisms may be predisposed to leptin resistance, with systemic consequences affecting energy metabolism, inflammation, and vascular integrity. Identifying such variants through genetic screening can improve risk stratification and guide personalized therapeutic interventions [26].
Given AMPK’s central role in these processes, therapeutic strategies that restore or enhance AMPK activity hold significant potential for improving cardiometabolic outcomes in this genetically susceptible population [26, 27].
**Therapeutic Strategies and Potential Interventions:**
Pharmacological activation of AMPK represents a key therapeutic approach. Agents such as metformin improve insulin sensitivity and promote fatty acid oxidation through AMPK activation, providing cardiovascular and metabolic benefits in individuals with leptin resistance. Natural compounds including resveratrol and berberine have also demonstrated AMPK-stimulating properties and may serve as adjunct therapies [27, 28].
Improving leptin sensitivity through lifestyle modifications can complement pharmacologic approaches. Regular physical activity and caloric restriction are known to enhance AMPK activation and leptin responsiveness. These interventions may counteract the effects of LEP variants by restoring energy balance and reducing cardiovascular risk [27].
Emerging therapies aimed at modulating leptin pathways may further expand treatment options. Leptin-mimetic agents or drugs that enhance leptin receptor sensitivity could restore physiological leptin signaling and indirectly promote AMPK activity, offering a targeted approach to managing obesity and associated cardiovascular complications [28].
**Implications:**
This study underscores the significance of genetic factors in modulating key metabolic pathways that influence cardiovascular risk. Understanding the role of LEP gene variants in disrupting leptin-AMPK interactions offers a mechanistic basis for the development of targeted, genotype-specific interventions. Integrating genetic screening into clinical practice could facilitate early detection of at-risk individuals and support precision medicine approaches in cardiometabolic care [28, 29].
Strategies that simultaneously address leptin resistance and AMPK dysfunction may yield synergistic benefits, reducing the burden of obesity-related cardiovascular disease and improving long-term patient outcomes [29].
Genetic variations in the LEP gene disrupt leptin signaling and impair AMPK activation, contributing to energy imbalance, chronic inflammation, and cardiovascular dysfunction. These variants promote obesity and its associated comorbidities through central and peripheral mechanisms affecting metabolism and vascular health. Therapeutic interventions that restore AMPK function or enhance leptin sensitivity represent promising approaches for mitigating cardiovascular risk in individuals with LEP-related obesity. Understanding the molecular interplay between leptin and AMPK may inform the development of precision therapies targeting obesity-induced cardiovascular disease [29, 30].
4. LEPR

**Genetic Variants in LEPR and Leptin Signaling Dysfunction:**
The LEPR gene encodes the leptin receptor, a transmembrane protein essential for mediating leptin’s regulatory effects on appetite and metabolism. Variations in LEPR disrupt receptor function and contribute to the pathophysiology of obesity. The single nucleotide polymorphism (SNP) rs1137101 (Q223R) is one of the most studied LEPR variants and is associated with altered receptor sensitivity and impaired leptin binding, often resulting in leptin resistance [31].
The leptin receptor plays a critical role in modulating AMP-activated protein kinase (AMPK) activity across central and peripheral tissues. Leptin’s effects on appetite suppression and metabolic regulation are partly mediated through AMPK signaling. When LEPR variants impair receptor function, the resulting changes in AMPK activation have downstream metabolic and cardiovascular implications [31, 32].
**AMPK Disruption from LEPR Variant-Induced Leptin Resistance:**
In the hypothalamus, leptin normally inhibits AMPK to suppress food intake and promote energy expenditure. Variants that impair leptin receptor function reduce this inhibition, resulting in increased hypothalamic AMPK activity. This overactivation can promote hyperphagia and disrupt energy balance, contributing to obesity [32].
In peripheral tissues such as liver and skeletal muscle, leptin activates AMPK to enhance fatty acid oxidation and glucose uptake. LEPR variants that impair receptor function lead to reduced AMPK activation, limiting lipid catabolism and promoting lipogenesis and insulin resistance. This dysfunctional metabolic profile exacerbates fat accumulation and impairs glycemic control [32, 33].
Leptin resistance, induced by LEPR polymorphisms, is also associated with dysregulated inflammatory signaling. Leptin enhances macrophage activation and cytokine release, and reduced AMPK activation diminishes the inhibition of pro-inflammatory pathways such as NF-κB. This shift favors a chronic low-grade inflammatory state, which is implicated in the pathogenesis of cardiovascular disease [33].
**Cardiovascular Consequences of Impaired AMPK Signaling:**
Impaired leptin signaling due to LEPR variants contributes to obesity and insulin resistance—two critical drivers of cardiovascular disease. Insulin resistance disrupts glucose homeostasis, promotes endothelial dysfunction, and accelerates atherosclerosis. Reduced AMPK activation in this context exacerbates metabolic dysfunction and promotes vascular inflammation [33, 34].
AMPK plays a key role in maintaining endothelial health by stimulating nitric oxide (NO) production and reducing oxidative stress. Attenuated AMPK activity due to leptin receptor dysfunction compromises endothelial function, leading to impaired vasodilation and increased oxidative damage. These changes heighten the risk of atherosclerotic plaque formation and cardiovascular events [34].
Leptin receptor signaling also modulates sympathetic nervous system (SNS) activity. Dysregulated leptin signaling may enhance SNS tone, contributing to elevated blood pressure. In combination with impaired AMPK signaling, this can promote the development of hypertension and its associated cardiovascular complications [34, 35].
**Clinical Relevance:**
The interplay between LEPR gene variants and AMPK signaling provides a mechanistic link between genetic susceptibility, metabolic dysregulation, and cardiovascular disease. Individuals with dysfunctional leptin receptor signaling may exhibit resistance to leptin’s anorexigenic and metabolic actions, resulting in a phenotype marked by hyperphagia, fat accumulation, and systemic inflammation [35].
From a clinical perspective, understanding the impact of LEPR variants on AMPK activation can inform risk stratification, early diagnosis, and tailored therapeutic interventions. Genetic screening for LEPR polymorphisms may help identify individuals at heightened cardiometabolic risk who would benefit from targeted AMPK-enhancing strategies [35, 36].
**Therapeutic Strategies and Targeted Interventions:**
AMPK activation represents a promising therapeutic avenue for counteracting the metabolic effects of LEPR variants. Agents such as metformin, which promote AMPK activation, have demonstrated efficacy in improving insulin sensitivity, stimulating fatty acid oxidation, and reducing systemic inflammation [36].
Non-pharmacological strategies including regular physical activity and caloric restriction also enhance AMPK signaling and improve leptin sensitivity. These lifestyle interventions may mitigate the downstream metabolic and cardiovascular effects of LEPR variant-induced leptin resistance [36, 37].
Targeted therapies that enhance leptin receptor function or mimic leptin’s effects may also prove beneficial. Development of leptin receptor agonists or leptin sensitizers could help restore AMPK activation and rebalance energy homeostasis, offering a more personalized approach to managing obesity and its cardiovascular sequelae in genetically predisposed individuals [37].
**Implications:**
The metabolic and cardiovascular consequences of LEPR variants underscore the need for integrating genomic information into precision medicine strategies. By elucidating the molecular pathways linking leptin resistance to AMPK dysfunction and cardiovascular pathology, this research supports the development of genotype-guided interventions [37, 38].
These findings highlight the potential for dual-targeted therapies that address both leptin signaling and AMPK activation to improve patient outcomes. Future therapeutic strategies should focus on reversing leptin resistance and restoring energy balance in individuals with LEPR-related obesity to reduce long-term cardiovascular risk [38].
Genetic variations in the LEPR gene contribute to leptin resistance and disrupt AMPK signaling, leading to metabolic dysregulation, chronic inflammation, and cardiovascular disease. These variants impair central and peripheral leptin-mediated metabolic control, promoting weight gain, insulin resistance, endothelial dysfunction, and hypertension. Therapeutic strategies aimed at restoring AMPK activity and enhancing leptin receptor function hold promise for addressing the metabolic and cardiovascular consequences associated with LEPR gene polymorphisms [39, 40].
5. PCSK1

**Role of PCSK1 in Hormone Processing and Energy Regulation:**
The PCSK1 gene encodes the enzyme prohormone convertase 1 (PC1/3), which plays a critical role in the post-translational processing of several metabolic hormones. PC1/3 is essential for converting inactive prohormones into biologically active peptides, including pro-opiomelanocortin (POMC), insulin, and glucagon-like peptide-1 (GLP-1). These hormones are central to regulating appetite, energy balance, glucose homeostasis, and lipid metabolism [41].
Disruptions in PC1/3 function, often due to genetic variants in PCSK1, impair the maturation of these hormones and have downstream effects on AMP-activated protein kinase (AMPK) signaling. Given AMPK’s role as a cellular energy sensor, its dysregulation contributes to metabolic and cardiovascular dysfunction, particularly in the context of obesity [41, 42].
**Impact of PCSK1 Variants on AMPK Activation:**
Common PCSK1 polymorphisms such as rs6232, rs6234, and rs6235 have been associated with obesity and features of metabolic syndrome. These variants are linked to reduced enzymatic activity or stability of PC1/3, leading to inefficient processing of POMC and insulin [42].
In the hypothalamus, POMC-derived peptides, particularly α-melanocyte-stimulating hormone (α-MSH), act on melanocortin receptors to inhibit AMPK activity, suppress appetite, and increase energy expenditure. Impaired POMC processing resulting from PCSK1 variants reduces α-MSH availability, resulting in persistent hypothalamic AMPK activation, which increases appetite and promotes energy storage [42, 43].
Peripheral effects of PCSK1 variants include impaired insulin maturation. Insulin plays a crucial role in activating AMPK in skeletal muscle and adipose tissue, promoting fatty acid oxidation and glucose uptake. A reduction in functional insulin due to deficient PC1/3 activity leads to decreased AMPK activation, insulin resistance, and lipid accumulation [43].
**Mechanistic Links Between PCSK1 Dysfunction and Metabolic Dysregulation:**
The metabolic consequences of impaired PC1/3 activity are mediated by disrupted hormonal signaling, which directly and indirectly modulates AMPK activity. Reduced POMC processing attenuates melanocortin signaling, contributing to increased AMPK activation in the hypothalamus, enhanced appetite, and subsequent weight gain. Simultaneously, insulin deficiency or dysregulation reduces AMPK activity in peripheral tissues, favoring lipogenesis and inhibiting energy expenditure [43, 44].
AMPK also plays an anti-inflammatory role by suppressing nuclear factor-kappa B (NF-κB) signaling and inflammatory cytokine production. Individuals with PCSK1 variants exhibit reduced AMPK activation, which may amplify inflammation, particularly in the context of insulin resistance. This chronic inflammatory state further exacerbates cardiovascular risk [44].
**Cardiovascular Complications of Altered AMPK Signaling in PCSK1 Variants:**
PCSK1 variants contribute to the pathogenesis of cardiovascular disease through multiple interconnected mechanisms. Insulin resistance, driven by impaired hormone processing and reduced AMPK activity, compromises endothelial function, disrupts lipid metabolism, and increases blood glucose levels. These factors collectively accelerate atherosclerotic processes [44, 45].
Endothelial dysfunction is closely linked to impaired AMPK activation, as AMPK promotes nitric oxide (NO) synthesis, maintains vascular tone, and mitigates oxidative stress. In PCSK1 variant carriers, reduced AMPK signaling undermines endothelial integrity, elevating the risk of hypertension and plaque formation [45].
Dyslipidemia, another hallmark of AMPK dysregulation, is commonly observed in individuals with PCSK1 mutations. Reduced AMPK activity results in impaired lipid oxidation and increased triglyceride synthesis, promoting vascular inflammation and stiffness. These effects compound the risk of cardiovascular events such as myocardial infarction and stroke [45, 46].
**Clinical Relevance:**
The interaction between PCSK1 gene variants and AMPK signaling underscores a crucial mechanistic link between genetic susceptibility, hormonal dysregulation, and cardiovascular pathology. Individuals harboring deleterious PCSK1 polymorphisms may present with early-onset obesity, insulin resistance, and an increased burden of cardiometabolic risk [46].
Understanding the hormonal basis of AMPK impairment in these individuals is essential for identifying high-risk phenotypes, guiding clinical decision-making, and designing personalized interventions. Early recognition of PCSK1-related metabolic dysfunction may facilitate preemptive strategies to mitigate long-term cardiovascular outcomes [46, 47].
**Therapeutic Approaches and Targeted Interventions:**
AMPK activators represent a central strategy for addressing metabolic dysfunction associated with PCSK1 variants. Pharmacologic agents such as metformin and thiazolidinediones activate AMPK, enhance insulin sensitivity, promote lipid catabolism, and reduce inflammatory signaling. These agents may partially restore metabolic balance in individuals with impaired PC1/3 function [47].
In severe cases, hormone replacement or supplementation may be necessary to compensate for deficiencies in insulin or other PC1/3-processed peptides. This approach could improve downstream AMPK activation and mitigate some of the metabolic disruptions observed in patients with PCSK1 mutations [47, 48].
Lifestyle interventions, including structured exercise and dietary modification, are effective non-pharmacological strategies for enhancing AMPK activity. Regular physical activity increases AMP/ATP ratios, activating AMPK and improving mitochondrial function, lipid oxidation, and glycemic control. These strategies are especially important in patients with PCSK1 variants and obesity-related cardiometabolic risk [48].
**Implications:**
The findings related to PCSK1 variants and their impact on AMPK signaling support a precision medicine approach to obesity and cardiovascular disease. Screening for PCSK1 polymorphisms may enable early identification of individuals at risk for hormone processing defects and allow for stratified therapeutic planning [48, 49].
The therapeutic value of AMPK activation in this genetic context is particularly compelling, suggesting that interventions aimed at restoring AMPK signaling may improve both metabolic and vascular outcomes. Future research should explore the development of targeted therapies that correct PC1/3 deficiencies or modulate the hormonal regulators of AMPK [49].
Genetic variations in the PCSK1 gene impair the processing of critical metabolic hormones and disrupt AMPK signaling pathways involved in appetite regulation, energy metabolism, and inflammation. These disruptions contribute to obesity, insulin resistance, and an elevated risk of cardiovascular disease. Therapeutic strategies focused on activating AMPK and correcting hormone imbalances hold promise for mitigating the metabolic and cardiovascular consequences associated with PCSK1-related obesity [50].
6. PPARG

**Functional Role of PPARG and Its Interaction with AMPK:**
The PPARG (Peroxisome Proliferator-Activated Receptor Gamma) gene encodes the nuclear receptor PPAR-γ, a critical regulator of lipid metabolism, adipocyte differentiation, and insulin sensitivity. PPAR-γ modulates energy balance by governing adipogenesis and lipid storage. It also interacts closely with AMP-activated protein kinase (AMPK), a central cellular energy sensor that maintains metabolic homeostasis. This interaction is particularly relevant in obesity, where disruptions in energy and lipid metabolism have downstream cardiovascular implications [51].
Genetic variations in PPARG, including the common polymorphisms Pro12Ala (rs1801282) and C161T (rs3856806), influence the activity of PPAR-γ. The Pro12Ala variant has been associated with enhanced insulin sensitivity, while the C161T variant may alter inflammatory responses and lipid regulation. These changes can impact AMPK activity, thereby affecting multiple physiological pathways central to cardiovascular health [51, 52].
**Genetic Variants and Effects on AMPK Activation:**
PPAR-γ modulates AMPK activation through transcriptional control of genes involved in lipid metabolism and insulin signaling. Variants in PPARG can influence the extent and efficiency of these regulatory functions. Altered PPAR-γ activity affects adipocyte lipid storage capacity, which has downstream consequences for AMPK-mediated fatty acid oxidation and systemic energy expenditure [52].
PPAR-γ enhances insulin sensitivity by promoting glucose uptake and adipokine production, particularly adiponectin. Adiponectin is a key activator of AMPK in peripheral tissues. Variants that reduce PPAR-γ function may lower adiponectin levels, decrease AMPK activation, and disrupt insulin-mediated energy regulation, which contributes to impaired glucose and lipid metabolism [52, 53].
**Mechanistic Pathways Linking PPARG Variants and AMPK Signaling:**
PPAR-γ is a central regulator of adipogenesis, promoting lipid uptake and storage in adipocytes. Variants that diminish PPAR-γ activity impair lipid handling and promote ectopic lipid deposition in non-adipose tissues such as liver and muscle, a process that reduces AMPK activation and contributes to lipotoxicity [53].
In addition to metabolic effects, PPAR-γ influences inflammatory signaling. It suppresses pro-inflammatory cytokines through transcriptional repression of NF-κB and other inflammatory pathways. When PPARG variants impair this function, a pro-inflammatory state may ensue. Given that AMPK exerts anti-inflammatory effects by inhibiting NF-κB, a reduction in AMPK activation linked to PPAR-γ dysfunction can amplify chronic inflammation and predispose individuals to cardiometabolic disease [53, 54].
PPAR-γ also regulates adipokine secretion, including adiponectin. Decreased adiponectin due to impaired PPAR-γ function results in reduced AMPK activation, contributing to insulin resistance, increased lipid accumulation, and systemic inflammation—core features of obesity-related cardiovascular pathology [54].
**Cardiovascular Pathophysiology Associated with Altered AMPK Signaling:**
Variants in PPARG that impair receptor function are associated with insulin resistance and central obesity, two key risk factors for cardiovascular disease. Reduced AMPK activation, secondary to impaired PPAR-γ signaling, contributes to endothelial dysfunction, impaired glucose utilization, and increased vascular inflammation [54, 55].
Endothelial dysfunction is a precursor to atherosclerosis and hypertension. AMPK plays a protective role in endothelial cells by promoting nitric oxide (NO) production and suppressing oxidative stress. Impairment in this pathway, due to altered PPAR-γ and reduced adiponectin, can compromise vascular tone and accelerate plaque development [55].
Dyslipidemia frequently accompanies PPARG dysfunction. Reduced AMPK activity exacerbates this condition by limiting fatty acid oxidation and enhancing hepatic lipid synthesis. This dysregulation contributes to elevated serum triglyceride levels and low HDL cholesterol, fostering atherogenic lipid profiles that increase cardiovascular risk [55, 56].
**Clinical Relevance:**
The relationship between PPARG gene variants and AMPK signaling underscores a significant genetic contribution to metabolic dysfunction and cardiovascular disease. Individuals harboring deleterious PPARG polymorphisms may exhibit early-onset insulin resistance, visceral adiposity, and systemic inflammation, collectively heightening their risk for cardiovascular events [56].
Understanding these genetic interactions provides insight into patient-specific risk profiles. This knowledge can inform clinical screening strategies and guide the selection of targeted therapies that address the underlying molecular dysfunction in high-risk individuals [56, 57].
**Therapeutic Interventions and Strategies:**
Pharmacological activation of PPAR-γ has been explored using thiazolidinediones (TZDs), such as rosiglitazone and pioglitazone. These agents improve insulin sensitivity and elevate adiponectin levels, indirectly enhancing AMPK activation. Although effective, their use requires careful risk-benefit analysis due to potential adverse cardiovascular effects associated with some TZDs [57].
AMPK activators, including metformin, provide an alternative or adjunct therapeutic option. Metformin directly activates AMPK, improves glucose uptake, enhances fatty acid oxidation, and suppresses inflammatory pathways. This makes it a valuable agent for mitigating cardiovascular risk in individuals with PPARG-associated metabolic dysfunction [57, 58].
Lifestyle interventions that stimulate AMPK activation, including regular physical activity and caloric restriction, represent non-pharmacologic strategies with well-documented efficacy. These interventions improve insulin sensitivity, reduce inflammation, and normalize lipid profiles, offering sustainable benefits in patients with PPARG-related obesity [58].
**Implications:**
The influence of PPARG variants on AMPK signaling suggests opportunities for precision medicine in the management of metabolic and cardiovascular disease. Genetic screening for PPARG polymorphisms may identify individuals who could benefit from early intervention and tailored treatment regimens targeting both PPAR-γ and AMPK pathways [58, 59].
Future research should explore dual-acting agents or combination therapies that simultaneously activate PPAR-γ and AMPK to maximize metabolic benefits while minimizing adverse effects. These strategies hold promise for improving cardiometabolic outcomes in genetically susceptible populations [59].
Genetic variations in the PPARG gene disrupt key pathways regulating lipid metabolism, insulin sensitivity, and inflammation, primarily through modulation of AMPK signaling. These disruptions play a central role in the development of obesity, insulin resistance, and cardiovascular disease. Therapeutic strategies that enhance PPAR-γ function, increase adiponectin levels, or directly activate AMPK offer promising avenues for reducing the cardiometabolic burden associated with PPARG-related genetic susceptibility [59, 60].
7. BDNF

**Functional Role of BDNF and Its Interaction with AMPK:**
The BDNF (Brain-Derived Neurotrophic Factor) gene encodes a neurotrophin crucial for neuronal development, synaptic plasticity, and neuroprotection. Beyond its neurological functions, BDNF plays a regulatory role in systemic energy homeostasis, appetite control, and cardiovascular health. These effects are mediated, in part, through modulation of AMP-activated protein kinase (AMPK), a central metabolic sensor involved in energy balance, inflammation, and vascular function. Genetic polymorphisms in BDNF have been implicated in metabolic disorders and cardiovascular disease, particularly in the context of obesity [61].
**Genetic Variants of BDNF and Their Effects on AMPK Activation:**
One of the most extensively studied BDNF polymorphisms is the Val66Met (rs6265) variant, which is associated with impaired activity-dependent BDNF secretion. Individuals carrying the Met allele display reduced central and peripheral BDNF bioavailability, which can lead to metabolic dysregulation [61, 62].
BDNF influences hypothalamic AMPK signaling, contributing to appetite suppression and energy expenditure. The Val66Met variant, by reducing BDNF secretion, can diminish hypothalamic AMPK activation, promoting hyperphagia and weight gain. In peripheral tissues, BDNF supports AMPK-mediated pathways involved in glucose uptake and fatty acid oxidation. Impaired BDNF signaling may consequently result in reduced AMPK activation, contributing to dysregulated glucose metabolism, lipid accumulation, and insulin resistance [62].
**Mechanisms Linking BDNF Variants to Altered AMPK Signaling:**
In the hypothalamus, BDNF exerts anorexigenic effects by activating AMPK and enhancing satiety signals. Variants that decrease BDNF availability can impair these signals, leading to increased food intake and decreased energy expenditure. This disruption in energy homeostasis contributes to positive energy balance and weight gain [62, 63].
Peripheral metabolic consequences of reduced BDNF signaling include impaired insulin sensitivity and reduced glucose uptake in skeletal muscle and hepatic tissues, partly due to diminished AMPK activation. These changes favor the development of insulin resistance and metabolic syndrome, key contributors to cardiovascular morbidity [63].
BDNF also modulates inflammatory signaling. AMPK activation suppresses pro-inflammatory transcription factors such as NF-κB. Reduced BDNF expression associated with the Val66Met variant may blunt AMPK activation, thereby enhancing systemic inflammation and increasing the risk of atherosclerosis and other cardiovascular complications [63, 64].
**Cardiovascular Consequences of Altered BDNF-AMPK Signaling:**
Reduced AMPK activation due to impaired BDNF signaling is associated with multiple cardiovascular risk factors. Obesity and insulin resistance, both linked to diminished AMPK activity, promote endothelial dysfunction and arterial stiffness. AMPK normally enhances endothelial nitric oxide production and reduces oxidative stress.
Impairment of this protective mechanism compromises vascular tone and promotes early atherogenesis [64].
Inflammation driven by reduced AMPK activity fosters plaque formation and progression of atherosclerosis. Additionally, dyslipidemia arising from impaired fatty acid oxidation and lipid clearance contributes to elevated serum triglyceride levels and low high-density lipoprotein (HDL) cholesterol. These lipid abnormalities further exacerbate cardiovascular risk [64, 65].
Hypertension may also result from disrupted vascular homeostasis. The loss of AMPK-mediated vasoprotective effects, in conjunction with systemic inflammation and metabolic dysregulation, contributes to elevated blood pressure and increased cardiovascular burden [65].
**Clinical Relevance:**
The Val66Met polymorphism and other BDNF gene variants represent significant genetic contributors to metabolic and cardiovascular risk. These variants influence appetite regulation, glucose metabolism, and inflammatory tone by modulating AMPK signaling pathways. Identifying individuals with BDNF polymorphisms may assist in early risk stratification and guide therapeutic interventions that address the underlying molecular dysfunctions [65, 66].
Genetic screening for BDNF variants, in combination with metabolic profiling, could enhance the precision of cardiovascular disease prevention strategies, especially in populations with high obesity prevalence or a family history of cardiometabolic disorders [66].
**Therapeutic Interventions:**
Pharmacological strategies to mitigate the effects of impaired BDNF signaling include the use of AMPK activators such as metformin. These agents promote glucose uptake, enhance fatty acid oxidation, and suppress inflammation, thereby compensating for reduced AMPK activity linked to BDNF variants [66, 67].
Emerging compounds that mimic or enhance BDNF signaling may offer an alternative route for restoring metabolic and vascular homeostasis. However, their clinical applicability remains under investigation [67].
Lifestyle interventions provide accessible and effective options for AMPK activation. Physical activity increases both BDNF expression and AMPK activity, supporting energy metabolism and cardiovascular function. Nutritional strategies that activate AMPK—such as caloric restriction, intermittent fasting, and diets rich in omega-3 fatty acids—can further reduce cardiovascular risk in genetically susceptible individuals [67, 68].
**Implications:**
BDNF variants that impair AMPK signaling highlight a critical genetic interface between neurotrophic regulation and systemic metabolic health. These findings reinforce the importance of integrative therapeutic approaches that address neuroendocrine, metabolic, and inflammatory dimensions of cardiovascular disease [68].
Incorporating genotypic information into clinical decision-making could support individualized interventions targeting both BDNF and AMPK pathways. Future research should explore the long-term outcomes of such targeted interventions and the development of dual-modulatory agents that restore neuro-metabolic balance and vascular integrity [68, 69].
Genetic variations in the BDNF gene, particularly the Val66Met polymorphism, influence AMPK signaling pathways involved in energy regulation, inflammation, and vascular health. These disruptions contribute to obesity, insulin resistance, and cardiovascular disease. Therapeutic approaches that enhance AMPK activation and restore BDNF signaling may offer effective strategies for managing cardiometabolic risk in individuals with genetically mediated BDNF dysfunction [70].
8. SIM1

**Role of SIM1 in Neuroendocrine Regulation:**
The SIM1 (Single-Minded Homolog 1) gene encodes a transcription factor that plays a pivotal role in hypothalamic development and neuroendocrine regulation. SIM1 expression is concentrated in the paraventricular nucleus of the hypothalamus, where it regulates key neuropeptides and hormones involved in appetite, energy balance, and metabolic homeostasis. Variants in SIM1 have been linked to early-onset obesity and altered feeding behavior, likely through their downstream impact on AMP-activated protein kinase (AMPK) signaling, a central regulator of cellular energy status [71].
**Genetic Variants of SIM1 and Their Effects on AMPK Activation:**
Several SIM1 polymorphisms, including the single nucleotide polymorphism rs13026078, have been associated with increased body mass index (BMI) and disrupted appetite regulation. These variants affect the transcriptional control of neuropeptides that modulate AMPK signaling. SIM1 influences the expression of melanocortins, neuropeptide Y (NPY), and other central effectors of energy metabolism, many of which converge on AMPK activity in the hypothalamus [71, 72].
Disruption of SIM1-regulated pathways can result in decreased AMPK activation in energy-sensing neuronal circuits, contributing to increased food intake and decreased energy expenditure. This imbalance promotes adiposity and metabolic dysregulation. Beyond central effects, SIM1 also influences hormonal mediators such as leptin and insulin, which are known regulators of AMPK in peripheral tissues. Impaired SIM1 function may therefore blunt AMPK activity both centrally and systemically [72].
**Mechanisms Linking SIM1 Variants to Altered AMPK Signaling:**
In the hypothalamus, AMPK regulates appetite and energy output in response to hormonal and nutritional cues. SIM1 variants that impair transcriptional activity may reduce the expression of melanocortins, weakening satiety signaling and diminishing AMPK-mediated suppression of appetite. This dysregulation contributes to chronic positive energy balance and obesity [72, 73].
Peripherally, SIM1 affects hormonal outputs that influence AMPK activation. Disruptions in leptin and insulin signaling, resulting from impaired SIM1 function, lead to reduced AMPK activation in skeletal muscle, adipose tissue, and liver. The consequences include impaired glucose uptake, increased insulin resistance, and reduced fatty acid oxidation [73].
SIM1 also indirectly influences inflammation through AMPK-dependent mechanisms. Reduced AMPK activity enhances pro-inflammatory cytokine production via pathways such as NF-κB, contributing to chronic inflammation and metabolic dysfunction [73, 74].
**Cardiovascular Consequences of Disrupted SIM1-AMPK Signaling:**
Obesity, insulin resistance, and chronic inflammation—hallmarks of impaired SIM1 function—are major contributors to cardiovascular disease (CVD). Variants in SIM1 that reduce AMPK activation may accelerate these pathological processes [74].
AMPK normally supports endothelial health by promoting nitric oxide (NO) bioavailability and reducing oxidative stress. Reduced AMPK signaling compromises endothelial function, increasing the risk of hypertension, vascular stiffness, and atherosclerotic plaque formation. Impaired lipid metabolism due to defective AMPK activation contributes to dyslipidemia, another critical CVD risk factor [74, 75].
Chronic inflammation, exacerbated by low AMPK activity, further drives atherosclerosis. Increased secretion of pro-inflammatory cytokines and recruitment of macrophages in vascular tissue promote the development of plaque and vascular occlusion, elevating the risk of myocardial infarction and stroke [75].
**Clinical Relevance:**
Variants in SIM1 are clinically significant due to their role in energy balance and metabolic disease. Individuals carrying pathogenic variants may present with early-onset obesity, hyperphagia, or resistance to traditional weight loss interventions. The link between SIM1 dysfunction and impaired AMPK signaling also positions these individuals at elevated risk for cardiometabolic disorders, including hypertension, atherosclerosis, and type 2 diabetes [75, 76].
Incorporating SIM1 genotyping into clinical risk assessment could improve the stratification of patients at risk for obesity-related cardiovascular disease. It may also inform therapeutic decisions, particularly regarding the use of metabolic modulators or behavioral interventions [76].
**Therapeutic Interventions:**
Pharmacological strategies that enhance AMPK activity hold potential in mitigating the metabolic consequences of SIM1 dysfunction. Metformin and other AMPK activators improve glucose metabolism, promote lipid oxidation, and suppress inflammation, offering benefits in insulin-resistant and obese patients with SIM1 variants [76, 77].
Lifestyle interventions remain foundational. Exercise is a potent activator of both SIM1 expression and AMPK signaling. Structured physical activity programs may be especially beneficial for individuals with impaired central energy regulation. Nutritional approaches that include caloric moderation and macronutrient optimization—such as diets rich in polyunsaturated fats and low in refined sugars—can further enhance AMPK activity and metabolic resilience [77].
Emerging therapies targeting hypothalamic circuits and neuropeptide signaling may also hold promise. Investigational approaches that modulate SIM1 transcriptional activity or mimic downstream neuropeptide effects could eventually restore energy balance in genetically susceptible individuals [77, 78].
**Implications:**
Understanding the genetic influence of SIM1 on AMPK signaling provides a mechanistic link between central neuroendocrine dysfunction and systemic cardiometabolic disease. This insight underscores the importance of integrative strategies that address both central and peripheral metabolic regulation [78].
Future research should aim to elucidate the full spectrum of SIM1 variants and their functional effects on AMPK and related signaling pathways. Clinical trials assessing the impact of AMPK-activating therapies in patients stratified by SIM1 genotype may further refine personalized approaches to cardiometabolic disease management [78, 79].
Genetic variations in the SIM1 gene contribute to metabolic dysregulation by impairing AMPK signaling in both central and peripheral tissues. These disruptions affect appetite control, energy expenditure, glucose metabolism, and inflammation, collectively increasing cardiovascular disease risk. Therapeutic strategies that enhance AMPK activation, restore hormonal signaling, or promote neuroendocrine homeostasis may offer targeted benefits for individuals with SIM1-related metabolic phenotypes [80].
9. TBC1D1

**Overview of TBC1D1 in Metabolic Regulation:**
TBC1D1 (TBC1 Domain Family Member 1) encodes a Rab-GTPase-activating protein that plays a critical role in the regulation of glucose uptake and lipid metabolism, primarily in skeletal muscle and adipose tissue. Its activity is closely linked to insulin signaling and AMP-activated protein kinase (AMPK) pathways, which are essential for energy homeostasis. Variants in TBC1D1 have been associated with obesity, insulin resistance, and cardiovascular disease (CVD). This review explores how genetic variations in TBC1D1 impact AMPK activation, alter downstream metabolic signaling, and contribute to cardiovascular risk [81].
**Genetic Variants of TBC1D1 and Their Effect on AMPK Activation:**
Several single nucleotide polymorphisms (SNPs) have been identified in TBC1D1, including rs1530871, which has been linked to increased body mass index (BMI), obesity, and insulin resistance. These variants can disrupt the normal function of the TBC1D1 protein, particularly its role in facilitating glucose transporter type 4 (GLUT4) translocation and mediating insulin-stimulated glucose uptake [81, 82].
TBC1D1 interacts with AMPK by serving as a substrate that facilitates GLUT4 trafficking to the cell membrane. Impaired TBC1D1 function due to genetic variations may reduce AMPK activation or compromise its downstream effects, limiting glucose uptake and promoting hyperglycemia. Additionally, TBC1D1 influences lipid oxidation, a process regulated by AMPK. Variants that reduce TBC1D1 function may decrease AMPK activity, shifting the metabolic profile toward lipid accumulation and reduced fatty acid oxidation [82].
**Mechanisms Linking TBC1D1 Variants to Altered AMPK Signaling:**
TBC1D1 is essential for regulating energy metabolism in response to insulin and exercise-induced stimuli. One of its primary roles is to promote GLUT4 translocation in skeletal muscle, facilitating glucose entry into cells. Genetic variants that impair this mechanism disrupt glucose homeostasis and reduce AMPK-mediated energy regulation [82, 83].
Disrupted insulin signaling is another consequence of TBC1D1 variation. Reduced AMPK phosphorylation resulting from impaired upstream signals leads to compromised glucose metabolism, decreased energy expenditure, and increased adiposity. TBC1D1 also influences lipid metabolism by regulating AMPK activity. When TBC1D1 is dysfunctional, AMPK fails to effectively promote fatty acid oxidation, contributing to lipogenesis and the development of obesity [83].
Inflammatory processes are also indirectly affected. AMPK typically exerts anti-inflammatory effects via suppression of nuclear factor kappa B (NF-κB) and other pro-inflammatory pathways. Reduced AMPK signaling due to TBC1D1 variants can enhance the expression of cytokines such as TNF-α and IL-6, perpetuating a chronic low-grade inflammatory state [83, 84].
**Impact on Cardiovascular Disease through AMPK Dysregulation:**
Impaired glucose uptake and altered lipid handling due to TBC1D1 dysfunction contribute to the pathogenesis of obesity and metabolic syndrome—both strong risk factors for CVD. Hyperglycemia and insulin resistance, hallmarks of reduced AMPK activity, can promote endothelial dysfunction and vascular inflammation [84].
Chronic inflammation plays a pivotal role in the development of atherosclerosis. With diminished AMPK activity, pro-inflammatory signaling is enhanced, increasing vascular permeability and promoting monocyte adhesion to endothelial cells. These changes accelerate plaque formation and heighten the risk of myocardial infarction and stroke [84, 85].
Endothelial dysfunction is exacerbated by oxidative stress and reduced nitric oxide (NO) bioavailability. AMPK activation supports endothelial health by promoting endothelial nitric oxide synthase (eNOS) activity. Impaired AMPK signaling due to TBC1D1 variants undermines this protective mechanism, predisposing individuals to hypertension and vascular disease [85].
Dyslipidemia is another downstream effect. When lipid oxidation is impaired and lipogenesis is increased, serum triglyceride and LDL cholesterol levels rise, worsening cardiovascular risk [86].
**Clinical Relevance:**
TBC1D1 variants represent a genetic determinant of metabolic flexibility and cardiovascular risk. Individuals harboring pathogenic variants may be predisposed to insulin resistance, obesity, and CVD, even in the absence of overt metabolic disease. Identifying such variants can aid in early risk stratification, particularly in patients with a family history of metabolic syndrome or premature cardiovascular events [86, 87].
Testing for TBC1D1 mutations could be integrated into personalized medicine approaches to guide therapeutic decisions, especially when considering the use of AMPK-targeting interventions or insulin-sensitizing agents [87].
**Therapeutic Interventions:**
Pharmacological AMPK activators, such as metformin and AICAR, offer promising avenues for restoring energy homeostasis in patients with TBC1D1 dysfunction. These agents enhance glucose uptake, promote fatty acid oxidation, and suppress inflammation—addressing multiple pathogenic mechanisms simultaneously [87, 88].
Exercise is a potent physiological activator of both TBC1D1 and AMPK. Regular physical activity can improve GLUT4 translocation, restore insulin sensitivity, and mitigate the metabolic impact of TBC1D1 variants [88].
Nutritional strategies that enhance AMPK activity, such as diets high in omega-3 fatty acids and low in refined carbohydrates, may further support metabolic resilience. These interventions can help optimize glucose and lipid metabolism, reduce inflammation, and improve vascular function [88, 89].
**Implications:**
Understanding the role of TBC1D1 genetic variants in modulating AMPK signaling provides a mechanistic link between inherited metabolic susceptibility and cardiovascular outcomes. As the field of cardiometabolic genomics evolves, TBC1D1 could serve as a valuable biomarker for predicting therapeutic response to AMPK-targeted treatments [89].
Targeted lifestyle and pharmacological interventions informed by genetic profiling may enable more effective management of cardiometabolic disorders. This highlights the growing importance of integrating genetic data into preventive and therapeutic frameworks for cardiovascular disease [89, 90].
Genetic variations in TBC1D1 significantly influence AMPK signaling pathways by disrupting glucose uptake, lipid oxidation, and inflammatory regulation. These molecular disruptions increase susceptibility to obesity, insulin resistance, and cardiovascular disease. Interventions that restore AMPK function—whether through pharmacological agents, physical activity, or dietary modifications—may provide effective strategies for mitigating the cardiovascular consequences associated with TBC1D1 dysfunction [90].
10. ADRB3

**Overview of ADRB3 in Metabolic Regulation:**
The ADRB3 (Beta-3 Adrenergic Receptor) gene encodes a G-protein-coupled receptor that plays a central role in the regulation of lipolysis and thermogenesis within adipose tissue. As part of the sympathetic nervous system, the β3-adrenergic receptor facilitates energy expenditure through the activation of key metabolic pathways. Its interaction with AMP-activated protein kinase (AMPK), a master regulator of energy homeostasis, suggests that ADRB3 genetic variants may influence metabolic flexibility and cardiovascular outcomes. This section explores how genetic variations in ADRB3 alter AMPK activation, modulate downstream metabolic responses, and contribute to cardiovascular disease (CVD) pathogenesis [91].
**Genetic Variants of ADRB3 and Their Impact on AMPK Activation:**
The most extensively studied polymorphism in ADRB3 is the Cys64Arg (rs4994) variant, which results in a substitution of arginine for cysteine at position 64. This mutation has been linked to obesity, reduced resting metabolic rate, and diminished lipolytic response to catecholamines [91, 92].
The β3-adrenergic receptor activates cyclic adenosine monophosphate (cAMP) production and subsequently protein kinase A (PKA), which may enhance AMPK activity. AMPK activation stimulates fatty acid oxidation and mitochondrial biogenesis, both critical for maintaining energy balance. Variants that impair ADRB3 function may disrupt this signaling cascade, resulting in reduced AMPK activation and compromised lipid metabolism. These disruptions can also diminish insulin sensitivity by limiting AMPK’s role in glucose uptake and lipid clearance, thereby exacerbating metabolic dysfunction [92].
**Mechanisms Linking ADRB3 Variants to Altered AMPK Signaling:**
The ADRB3 receptor is predominantly expressed in brown and white adipose tissue and contributes to adaptive thermogenesis and lipid mobilization. Variants that impair receptor activation compromise AMPK-mediated processes essential for energy regulation [92, 93].
In brown adipose tissue, impaired thermogenesis due to reduced β3-adrenergic signaling can decrease AMPK activity and blunt fatty acid oxidation. In white adipose tissue, reduced lipolysis diminishes free fatty acid availability for energy production, limiting AMPK activation. These alterations favor fat accumulation and promote insulin resistance [93].
AMPK also exerts anti-inflammatory effects, including suppression of nuclear factor kappa B (NF-κB) signaling. A decline in AMPK activity caused by dysfunctional ADRB3 signaling may increase levels of pro-inflammatory cytokines such as TNF-α and IL-6. These cytokines impair insulin signaling and further disrupt glucose metabolism, establishing a feedback loop that worsens metabolic homeostasis [93, 94].
**Impact on Cardiovascular Disease via AMPK Dysregulation:**
Reduced AMPK activity due to impaired ADRB3 signaling contributes to multiple cardiovascular risk factors. Obesity and metabolic syndrome are more prevalent in individuals carrying the Cys64Arg variant, reflecting impaired energy expenditure and increased adiposity. This metabolic phenotype is closely associated with insulin resistance, endothelial dysfunction, and systemic inflammation—all of which are major contributors to cardiovascular pathology [94].
Chronic inflammation plays a central role in atherogenesis. Decreased AMPK signaling increases the expression of adhesion molecules and cytokines, promoting monocyte recruitment and plaque formation. In addition, reduced nitric oxide (NO) production due to insufficient AMPK activation compromises endothelial function and vascular reactivity, raising the risk of hypertension and atherosclerotic events [94, 95].
Dyslipidemia also emerges from disrupted AMPK function. Inadequate lipid oxidation and continued lipogenesis lead to elevated plasma triglycerides and reduced HDL cholesterol, enhancing the likelihood of coronary artery disease and stroke [95].
**Clinical Relevance:**
The presence of ADRB3 variants, particularly Cys64Arg, may serve as a genetic marker for increased susceptibility to obesity, insulin resistance, and cardiovascular disease. Understanding the functional consequences of these variants can inform early screening and risk stratification, especially in patients presenting with unexplained metabolic abnormalities [95, 96].
Genotyping for ADRB3 polymorphisms may provide predictive value in personalized treatment approaches, identifying individuals who may benefit most from AMPK-targeted therapies or β3-adrenergic receptor modulators [96].
**Therapeutic Strategies and Potential Interventions:**
Pharmacological strategies to counteract the effects of ADRB3 variants include the use of AMPK activators, such as metformin or AICAR, which promote fatty acid oxidation, improve insulin sensitivity, and suppress inflammatory signaling. These agents may restore metabolic balance in individuals with impaired β3-adrenergic signaling [96, 97].
Targeted β3-adrenergic receptor agonists offer another avenue for therapeutic intervention, particularly in enhancing energy expenditure and lipolysis. While some agents are under investigation, further clinical studies are needed to evaluate their efficacy in genetically susceptible populations [97].
Lifestyle interventions remain foundational. Exercise is a well-established activator of AMPK and can improve both glucose metabolism and mitochondrial function. Structured physical activity can offset the metabolic consequences of ADRB3 dysfunction by promoting energy expenditure and reducing adiposity [97, 98].
Dietary modifications may further support AMPK activation. Diets enriched in omega-3 fatty acids and low in saturated fats and refined sugars can improve lipid profiles, enhance mitochondrial function, and support metabolic flexibility. These strategies may be particularly beneficial in individuals with known ADRB3 polymorphisms [98].
**Implications:**
The interaction between ADRB3 gene variants and AMPK signaling underscores the complex genetic and molecular determinants of metabolic and cardiovascular disease. Elucidating these mechanisms provides a path toward precision medicine, enabling the development of individualized interventions based on genetic risk [98, 99].
Recognition of the ADRB3-AMPK axis as a modifiable target highlights opportunities for pharmacologic and behavioral strategies to mitigate cardiometabolic risk. These insights have implications for public health strategies aimed at reducing the burden of obesity-related cardiovascular disease through genetic screening and tailored therapies [99].
Genetic variations in the ADRB3 gene, particularly the Cys64Arg polymorphism, influence AMPK signaling by impairing lipolysis, energy expenditure, and anti-inflammatory responses. These disruptions contribute to the pathogenesis of obesity, insulin resistance, and cardiovascular disease. Targeted therapeutic strategies— including AMPK activation, β3-adrenergic receptor modulation, and lifestyle interventions—offer potential for mitigating the cardiometabolic consequences of ADRB3 dysfunction. Continued research into this genetic-metabolic axis will be essential for advancing personalized approaches to cardiovascular risk management [100].
11. UCP1

**Overview of UCP1 in Metabolic Regulation:**
The UCP1 (Uncoupling Protein 1) gene encodes a mitochondrial inner membrane protein predominantly expressed in brown adipose tissue (BAT), where it plays a central role in non-shivering thermogenesis. UCP1 dissipates the proton gradient generated during oxidative phosphorylation, releasing energy as heat rather than synthesizing ATP. This thermogenic process contributes significantly to total energy expenditure and is closely modulated by AMP-activated protein kinase (AMPK), a master regulator of energy homeostasis. Understanding the molecular relationship between UCP1 genetic variations and AMPK activation provides valuable insight into the pathophysiology of obesity and cardiovascular disease (CVD) [101].
**Genetic Variants of UCP1 and Their Impact on AMPK Activation:**
Several single nucleotide polymorphisms (SNPs) in UCP1, such as rs1800592, have been associated with alterations in energy metabolism, body mass index (BMI), and predisposition to metabolic disorders. These genetic variants may reduce the expression or activity of UCP1, thereby impairing thermogenesis [101, 102].
AMPK is activated in response to decreased intracellular ATP levels and orchestrates compensatory energy-generating pathways, including glucose uptake and fatty acid oxidation. Impaired UCP1 function due to genetic variants may reduce thermogenic energy expenditure, lowering cellular energy turnover and attenuating AMPK activation. This disruption can compromise the cell’s ability to maintain energy balance, predisposing individuals to obesity and metabolic dysfunction [102].
**Mechanistic Links Between UCP1 Variants and AMPK Signaling:**
UCP1-mediated thermogenesis serves as a key mechanism for adaptive energy dissipation in response to cold exposure and nutrient excess. Variants that diminish UCP1 expression or functionality can blunt this thermogenic capacity, decreasing the energetic demand that normally activates AMPK [102, 103].
Reduced substrate oxidation due to impaired UCP1 function may limit the generation of AMP, leading to insufficient AMPK activation. This limitation affects downstream metabolic targets, including enzymes involved in lipid oxidation and glucose transport. Since AMPK also enhances insulin sensitivity through upregulation of GLUT4-mediated glucose uptake and inhibition of hepatic gluconeogenesis, its reduced activation exacerbates insulin resistance [103].
Impaired AMPK signaling in individuals with UCP1 variants may also hinder the mobilization and oxidation of fatty acids, contributing to increased adiposity and altered lipid profiles. These metabolic consequences collectively disrupt energy homeostasis and increase cardiovascular risk [103, 104].
**Contribution to Cardiovascular Disease:**
Genetic variations in UCP1 that result in reduced AMPK activity are associated with increased incidence of obesity and metabolic syndrome, both major risk factors for cardiovascular disease.
Chronic inflammation is a common feature of obesity and plays a central role in atherosclerosis. AMPK negatively regulates pro-inflammatory pathways such as nuclear factor-kappa B (NF-κB). Diminished AMPK signaling due to impaired UCP1 activity may promote overproduction of inflammatory cytokines, facilitating vascular inflammation and plaque formation [104].
Endothelial dysfunction is a critical early event in the development of hypertension and atherosclerosis. AMPK activation stimulates endothelial nitric oxide synthase (eNOS), enhancing nitric oxide (NO) bioavailability and promoting vasodilation. Reduced AMPK activity secondary to UCP1 dysfunction may compromise NO production, increasing vascular tone and oxidative stress [104, 105].
Alterations in lipid metabolism due to inadequate AMPK activation further increase cardiovascular risk. Individuals with UCP1 variants may exhibit dyslipidemia characterized by elevated triglycerides and reduced HDL cholesterol, promoting atherogenesis and vascular complications [105].
**Clinical Relevance:**
UCP1 genetic variants represent a clinically significant factor in the regulation of thermogenesis and metabolic health. Understanding the effect of these variants on AMPK signaling may support the early identification of individuals at risk for metabolic syndrome and cardiovascular disease. Given the rising prevalence of obesity, the UCP1-AMPK axis offers a potential target for metabolic screening and personalized intervention [105, 106].
The presence of SNPs such as rs1800592 may serve as biomarkers for predicting metabolic flexibility, responsiveness to lifestyle interventions, or pharmacologic treatments targeting energy expenditure and insulin sensitivity [106].
**Implications:**
Therapeutic strategies that enhance UCP1 expression or mimic its effects may restore AMPK activity and improve metabolic outcomes in genetically predisposed individuals. AMPK activators, such as metformin or AICAR, can directly enhance fatty acid oxidation and improve insulin sensitivity, potentially counteracting the metabolic impairments linked to UCP1 dysfunction [106, 107].
Physical activity is a potent natural activator of AMPK and has been shown to increase UCP1 expression in BAT. Regular exercise may help ameliorate the adverse effects of UCP1 polymorphisms by improving thermogenesis, glucose metabolism, and endothelial function [107].
Nutritional approaches aimed at stimulating thermogenesis and AMPK activation—such as diets high in polyunsaturated fatty acids, low in simple sugars, and incorporating cold exposure or thermogenic nutrients— could offer additional benefit in managing cardiometabolic risk in individuals with UCP1 variants [107, 108].
Genetic variations in the UCP1 gene modulate thermogenesis and energy metabolism via the AMPK signaling pathway. Impaired UCP1 function can diminish AMPK activation, leading to reduced energy expenditure, insulin resistance, inflammation, and endothelial dysfunction. These changes increase the risk of obesity and cardiovascular disease. Integrative therapeutic approaches that enhance UCP1 function, activate AMPK, and promote metabolic resilience through lifestyle modification may reduce the burden of cardiometabolic disease associated with UCP1 genetic polymorphisms [109, 110].
12. SH2B1

**Overview of SH2B1 in Metabolic Regulation:**
The SH2B1 (SH2B Adaptor Protein 1) gene encodes a signaling adaptor protein that plays a critical role in regulating metabolic processes through its interactions with key hormonal pathways, including insulin and leptin signaling. These pathways intersect with AMP-activated protein kinase (AMPK), a master regulator of cellular energy homeostasis. Genetic variants in SH2B1 have been linked to obesity, insulin resistance, and other metabolic abnormalities. This investigation explores how SH2B1 variants modulate AMPK activation, their impact on energy balance and inflammation, and the downstream consequences for cardiovascular health [111].
**Genetic Variants in SH2B1 and Their Effects on AMPK Activation:**
Several SH2B1 single nucleotide polymorphisms (SNPs), such as rs7498665, have been associated with increased body mass index (BMI) and susceptibility to obesity. These variants are believed to disrupt the normal function of SH2B1, impairing its ability to facilitate signal transduction in insulin and leptin pathways [111, 112].
SH2B1 enhances insulin sensitivity, and insulin signaling is known to promote AMPK activation under energy-depleted conditions. Genetic variants that attenuate SH2B1 function may reduce insulin signaling efficacy, resulting in insufficient AMPK activation. Similarly, SH2B1 mediates leptin receptor signaling, which regulates appetite and energy expenditure. Disruption in this axis due to genetic variation may compromise AMPK activation, limiting its ability to coordinate metabolic homeostasis [112].
**Mechanisms Linking SH2B1 Variants to Altered AMPK Signaling:**
AMPK plays a central role in balancing energy intake and expenditure. In tissues such as skeletal muscle, AMPK promotes glucose uptake and fatty acid oxidation. SH2B1 variants that impair insulin and leptin signaling may result in lower AMPK activation, diminishing glucose transport and reducing mitochondrial fat oxidation. This leads to energy surplus and adiposity [112, 113].
Inflammation is another critical domain influenced by the SH2B1–AMPK interaction. AMPK negatively regulates the expression of pro-inflammatory cytokines. Genetic variants in SH2B1 may disturb this anti-inflammatory pathway, resulting in elevated cytokine production. Chronic low-grade inflammation is a well-established driver of metabolic dysfunction and cardiovascular disease, suggesting that impaired SH2B1 function contributes to these pathologies by undermining AMPK-mediated immune regulation [113].
**Cardiovascular Consequences of SH2B1-Mediated AMPK Dysregulation:**
Disruption of SH2B1 function through genetic variation contributes to obesity, insulin resistance, and systemic inflammation, which are key components of metabolic syndrome and primary risk factors for cardiovascular disease. These metabolic abnormalities, compounded by impaired AMPK activity, create a pro-atherogenic and pro-inflammatory environment [113, 114].
Reduced AMPK signaling is associated with increased production of pro-inflammatory cytokines such as IL-6 and TNF-α. These mediators accelerate the progression of atherosclerosis and contribute to vascular injury.
Endothelial dysfunction is also exacerbated by reduced AMPK activation, which limits nitric oxide (NO) synthesis and promotes oxidative stress. SH2B1 variants that inhibit AMPK activation thereby compromise vascular integrity and elevate the risk of hypertension and ischemic events [114].
Dyslipidemia is frequently observed in individuals with impaired AMPK activity, marked by elevated serum triglycerides and reduced high-density lipoprotein (HDL) cholesterol. SH2B1 variants may promote this atherogenic lipid profile by altering AMPK’s regulatory effects on hepatic lipid metabolism [114, 115].
**Clinical Relevance:**
The identification of SH2B1 genetic variants in individuals with obesity or cardiometabolic disorders may offer diagnostic and prognostic insights. These variants serve as potential genetic markers for predisposition to insulin resistance, dyslipidemia, and cardiovascular complications. Understanding the link between SH2B1 and AMPK activation provides an opportunity to personalize prevention and treatment strategies in at-risk populations [115].
Clinicians may consider genetic screening for SH2B1 polymorphisms to stratify cardiovascular risk or guide the choice of therapeutic interventions aimed at enhancing AMPK signaling. Patients with these variants may require more aggressive lifestyle modification or pharmacologic interventions to prevent progression to cardiovascular disease [115, 116].
**Implications for Therapy and Intervention:**
Targeted therapeutic approaches can be developed to counteract the metabolic impact of SH2B1 variants. Pharmacological AMPK activators such as metformin and thiazolidinediones have shown efficacy in improving insulin sensitivity and reducing systemic inflammation, potentially mitigating the metabolic consequences of SH2B1 dysfunction [116].
Non-pharmacological strategies also play a critical role. Structured physical activity is a potent activator of AMPK and improves mitochondrial function, glucose uptake, and lipid oxidation. Exercise may serve as an effective intervention for individuals with SH2B1 variants by partially restoring impaired AMPK signaling [116, 117].
Nutritional interventions that favor AMPK activation can further support metabolic health. Diets rich in polyunsaturated fatty acids, fiber, and micronutrients, along with the avoidance of refined sugars and trans fats, may enhance AMPK responsiveness and improve lipid profiles in genetically susceptible individuals [118].
Genetic variations in SH2B1 influence critical metabolic and inflammatory pathways through modulation of AMPK signaling. These variants contribute to the development of obesity, insulin resistance, and chronic inflammation, increasing cardiovascular disease risk. Recognition of these gene–pathway interactions enables more precise identification of at-risk individuals and supports the design of targeted therapeutic strategies.
Enhancing SH2B1 function, restoring AMPK activity, and encouraging lifestyle changes represent promising avenues to reduce cardiometabolic risk in affected populations [119, 120].

## Discussion

### Genetic Variations in Obesity-Related Genes and Their Metabolic Significance

This study provides key insights into the impact of genetic variations in obesity-related genes on the AMP-activated protein kinase (AMPK) signaling pathway. The results highlight the intricate relationship between genetic predisposition, energy metabolism, and cardiovascular health. Genes such as FTO, MC4R, LEP, and LEPR play central roles in regulating appetite, satiety, and energy expenditure. Variants in these genes appear to modulate susceptibility to obesity by altering neuroendocrine signaling mechanisms that govern caloric intake and storage.

Understanding the contribution of these polymorphisms to metabolic dysfunction is essential for interpreting individual variability in obesity development and progression. In particular, the dysregulation of leptin signaling pathways due to altered LEP and LEPR activity underscores the potential of targeting this axis for therapeutic modulation of appetite and energy balance.

### AMPK Signaling and Therapeutic Opportunities

The study underscores the centrality of AMPK as a molecular hub in energy regulation. AMPK activation promotes glucose uptake, fatty acid oxidation, and mitochondrial biogenesis, supporting overall metabolic homeostasis. Genetic alterations that impair AMPK signaling reduce its protective metabolic functions, predisposing individuals to insulin resistance, lipid accumulation, and metabolic syndrome.

Given these findings, therapeutic strategies that enhance AMPK activation may be particularly advantageous for individuals harboring obesity-related gene variants. Lifestyle interventions such as regular physical activity and calorie restriction, alongside pharmacological agents including metformin and AICAR, offer viable approaches to reactivating AMPK pathways. Tailoring these interventions based on individual genetic profiles may enable precision strategies to counteract metabolic dysfunction in high-risk populations.

### Inflammation, Endothelial Dysfunction, and Cardiovascular Risk

The downstream consequences of impaired AMPK signaling extend beyond metabolic disturbances. Reduced AMPK activity contributes to vascular inflammation, endothelial dysfunction, and diminished nitric oxide bioavailability—key mechanisms implicated in the pathogenesis of cardiovascular disease. Genetic variants affecting AMPK regulation may thereby influence a pro-inflammatory state that accelerates the development of atherosclerosis, hypertension, and other cardiovascular complications in obese individuals.

Addressing inflammation through targeted therapies, such as anti-inflammatory agents or AMPK activators, may offer dual benefits in restoring vascular integrity and improving metabolic resilience. Nutritional interventions designed to reduce systemic inflammation, such as increased intake of polyphenols or omega-3 fatty acids, may also serve as adjunctive strategies in cardiovascular risk management.

### Clinical Relevance

These findings underscore the need for integrating genetic screening into the clinical assessment of patients with obesity. Identification of individuals with specific risk alleles may inform early interventions to prevent metabolic derangements and cardiovascular complications. AMPK activation emerges as a promising therapeutic target, with potential for both preventive and corrective strategies in genetically predisposed populations. Personalized treatment approaches based on genetic profiles can enhance efficacy and minimize adverse effects, particularly in the management of obesity, type 2 diabetes, and cardiovascular comorbidities.

### Implications

This study supports the development of genotype-guided treatment protocols that address both metabolic and vascular consequences of obesity. Clinical implementation of such approaches will require the incorporation of genetic testing into routine practice, alongside validated guidelines for interpreting and acting upon genotypic data. The findings also advocate for public health initiatives promoting early lifestyle interventions in individuals with known genetic susceptibility, with the goal of reducing the long-term burden of obesity-related diseases.

### Future Research Directions

Further research is needed to delineate the precise molecular mechanisms through which obesity-related genetic variants disrupt AMPK signaling. Studies exploring gene-environment interactions—particularly the modifying effects of diet, exercise, and pharmacologic agents—will be vital for optimizing personalized care. Large-scale, longitudinal studies are warranted to evaluate the long-term cardiovascular outcomes associated with these variants and to validate therapeutic approaches tailored to genetic risk profiles.

This investigation highlights the pivotal role of genetic factors in modulating AMPK signaling and influencing metabolic and cardiovascular outcomes in obesity. A deeper understanding of these genetic influences offers new avenues for personalized interventions aimed at restoring metabolic balance and reducing cardiovascular risk. Addressing the genetic underpinnings of obesity is an essential step toward advancing precision medicine and improving health outcomes in populations worldwide.

## Conclusion

This investigation elucidates the intricate relationships between genetic variations in key obesity-related genes—FTO, MC4R, LEP, LEPR, PCSK1, PPARG, BDNF, SIM1, TBC1D1, ADRB3, UCP1, and SH2B1—and the AMPK signaling pathway. Each gene plays a pivotal role in the regulation of energy homeostasis and metabolic processes, and their genetic variations can significantly affect AMPK activation and its downstream effects.

The findings reveal that alterations in these genes can lead to impaired AMPK signaling, resulting in decreased energy expenditure, increased lipid accumulation, and disrupted glucose metabolism. For instance, variations in FTO and MC4R contribute to dysregulated appetite and energy balance, while LEP and LEPR variations affect leptin signaling pathways that regulate energy homeostasis. Similarly, genetic alterations in PCSK1 and PPARG impact insulin sensitivity and lipid metabolism, which are closely tied to AMPK activity.

Moreover, genes such as UCP1 and SH2B1 illustrate how thermogenic and adaptor proteins interact with AMPK to modulate energy expenditure and inflammation. Collectively, these genetic variations contribute to a heightened inflammatory state, increased oxidative stress, and endothelial dysfunction, all of which are implicated in the pathogenesis of cardiovascular disease.

The interplay between genetic variations in obesity-related genes and the AMPK signaling pathway underscores the complexity of metabolic regulation in obesity. These interactions not only highlight the potential mechanisms by which obesity contributes to cardiovascular pathology but also point to therapeutic avenues targeting AMPK activation and gene-specific interventions that may ameliorate metabolic dysregulation and reduce cardiovascular risks in individuals with obesity. Further research is warranted to elucidate the precise molecular mechanisms involved and to explore personalized strategies for managing obesity-related cardiovascular disease.

## Abbreviations

AMPK: AMP-activated protein kinase
CVD: Cardiovascular disease
FTO: Fat Mass and Obesity-Associated gene
MC4R: Melanocortin 4 Receptor
LEP: Leptin gene
LEPR: Leptin Receptor
PCSK1: Proprotein Convertase Subtilisin/Kexin Type 1
PPARG: Peroxisome Proliferator-Activated Receptor Gamma
BDNF: Brain-Derived Neurotrophic Factor
SIM1: Single-minded homolog 1
TBC1D1: TBC1 Domain Family Member 1
ADRB3: Beta-3 Adrenergic Receptor
UCP1: Uncoupling Protein 1
SH2B1: SH2B Adaptor Protein 1
PRISMA: Preferred Reporting Items for Systematic Reviews and Meta-Analyses
SNP: Single Nucleotide Polymorphism
BMI: Body Mass Index
PGC-1α: Peroxisome proliferator-activated receptor gamma coactivator 1-alpha
HDL: High-Density Lipoprotein
TNF-α: Tumor Necrosis Factor Alpha
IL-6: Interleukin-6
NO: Nitric Oxide
GLUT4: Glucose Transporter Type 4

## Declarations

### Ethics declarations

**Ethics approval and consent to participate**

Not applicable.

### Consent for publication

Not applicable.

### Data Availability statement

All data generated or analyzed during this study are included in this article.

## Acknowledgements

“Not applicable”.

## Author’s Information

1. **Ifrah Siddiqui (IS)\*** is the author of the study and contributed to its conceptualization, design, and methodology, as well as the literature search and referencing. She was responsible for writing, editing, and revising the manuscript, as well as delineating the findings, results, conclusions, implications, and all other aspects of the study. IS conducted data extraction and analysis, critically evaluated every aspect of the study, ensured adherence to relevant PRISMA guidelines, and addressed study limitations and references. Additionally, she created PRISMA Flow Diagram.

She investigated how genetic variations in key obesity-related genes — FTO, MC4R, LEP, LEPR, PCSK1, PPARG, BDNF, SIM1, TBC1D1, ADRB3, UCP1, and SH2B1 — influence the AMPK signaling pathway and its downstream effects on energy homeostasis, inflammation, and cardiovascular disease risk in the context of obesity.
Ifrah Siddiqui (IS)* holds a Bachelor’s Degree with a focus on Psychology from the University of Karachi, Pakistan. Currently, she is undertaking training/course in Health & Medicine from Harvard Medical School. She has a passion for investigating the molecular mechanisms underlying disease pathogenesis and psychological aspects of various diseases.
Corresponding author: IS
Correspondence to <u>Ifrah Siddiqui</u>
2. **Nabeel Ahmad Khan (NAK)** is the co-author of the study and contributed to the writing, editing and revision. He contributed to the background, results, discussion and conclusion sections of the study along with working on the findings, interpretation of the data and references.

He contributed to the investigation of FTO, MC4R, PPARG, and BDNF genes.
Nabeel Ahmad Khan (NAK) holds a Doctor of Pharmacy (Pharm.D.) degree from Dow University of Health Sciences, Karachi, Pakistan, completed in 2015, and a Master’s in Multidisciplinary Biomedical Sciences with a concentration in pharmacology from the University of Alabama at Birmingham, completed in 2022. His research interests include exploring the causal link between environmental factors and disease pathogenesis. Nabeel is dedicated to understanding how environmental influences contribute to the development and progression of various diseases, aiming to identify new therapeutic targets and strategies. Through his work, he seeks to advance the field of biomedical sciences by uncovering critical insights that can lead to improved disease prevention and treatment.
3. **Zille Haider** is the co-author of the study and contributed to the writing, editing and revision. He contributed to the background, results and conclusion sections of the study.

He contributed to the investigation of LEP, LEPR, and SH2B1 genes.
Zille Haider holds an MBBS degree from Fatima Memorial Hospital College of Medicine & Dentistry, Pakistan. His academic and research interests focus on the intersection of molecular genetics, metabolic disorders, and cardiovascular disease, with a particular emphasis on obesity-related mechanisms.
*The work and* contributions *of everyone have been described in detail, the order is randomized and the numbering is just for referencing purpose*.

